# Effectiveness of Primary and Booster COVID-19 mRNA Vaccination against Omicron Variant SARS-CoV-2 Infection in People with a Prior SARS-CoV-2 Infection

**DOI:** 10.1101/2022.04.19.22274056

**Authors:** Margaret L. Lind, Alexander James Robertson, Julio Silva, Frederick Warner, Andreas C. Coppi, Nathan Price, Chelsea Duckwall, Peri Sosensky, Erendira C. Di Giuseppe, Ryan Borg, Mariam O Fofana, Otavio T. Ranzani, Natalie E. Dean, Jason R. Andrews, Julio Croda, Akiko Iwasaki, Derek A.T. Cummings, Albert I. Ko, Matt DT Hitchings, Wade L. Schulz

## Abstract

**Background:** The benefit of vaccination in people who experienced a prior SARS-CoV-2 infection remains unclear.

**Objective:** To estimate the effectiveness of primary (two-dose) and booster (third dose) vaccination against Omicron infection among people with a prior documented infection.

**Design:** Test-negative case-control study.

**Setting:** Yale New Haven Health System facilities.

**Participants:** Vaccine eligible people who received SARS-CoV-2 RT-PCR testing between November 1, 2021, and January 31, 2022.

**Measurements:** We conducted two analyses, each with an outcome of Omicron BA.1 infection (S-gene target failure defined) and each stratified by prior SARS-CoV-2 infection status. We estimated the effectiveness of primary and booster vaccination. To test whether booster vaccination reduced the risk of infection beyond that of the primary series, we compared the odds among boosted and booster eligible people.

**Results:** Overall, 10,676 cases and 119,397 controls were included (6.1% and 7.8% occurred following a prior infection, respectively). The effectiveness of primary vaccination 14-149 days after 2^nd^ dose was 36.1% (CI, 7.1% to 56.1%) for people with and 28.5% (CI, 20.0% to 36.2%) without prior infection. The odds ratio comparing boosted and booster eligible people with prior infection was 0.83 (CI, 0.56 to 1.23), whereas the odds ratio comparing boosted and booster eligible people without prior infection was 0.51 (CI, 0.46 to 0.56).

**Limitations:** Misclassification, residual confounding, reliance on TaqPath assay analyzed samples.

**Conclusion:** While primary vaccination provided protection against BA.1 infection among people with and without prior infection, booster vaccination was only associated with additional protection in people without prior infection. These findings support primary vaccination in people regardless of prior infection status but suggest that infection history should be considered when evaluating the need for booster vaccination.

**Primary Funding Source:** Beatrice Kleinberg Neuwirth and Sendas Family Funds, Merck and Co through their Merck Investigator Studies Program, and the Yale Schools of Public Health and Medicine.

## Introduction

Although COVID-19 vaccines provide lower levels of protection against the B.1.1.529 (Omicron) than the B.1.617.2 (Delta) variant of SARS-CoV-2, current evidence indicates that primary and booster (third) vaccination significantly reduces the risk of Omicron related outcomes in the general population.(1–5) However, the benefit of vaccination in people with a prior SARS-CoV-2 infection remains uncertain. Previous studies, conducted prior to the Omicron epidemic wave, found that primary vaccination (two doses) afforded protection against reinfection beyond that provided by a prior infection(6–9) and that a booster dose significantly increase such protection.(10) In contrast, Shrestha et al. found that primary vaccination did not provide additional protection (Hazard Ratio, 0.77; 95% CI, 0.53-1.12) against SARS-CoV-2 re-infection among previously infected people during the first month of the Omicron wave.(11) Furthermore, evidence is lacking for the additional benefit of booster vaccination against Omicron infection in individuals with a prior infection, which is needed to inform vaccination policies for this sub-population.

In this study, we analyzed data from a large cohort of people from the Yale New Haven Health system who underwent molecular testing for S-gene target failure (SGTF) to evaluate the benefit of primary series and booster doses in the context of the Omicron wave. Specifically, we estimated the effectiveness of primary and booster vaccination against Omicron (lineage BA.1) infection among people with and without a documented prior SARS-CoV-2 infection. We also examined whether booster vaccination reduced the risk of Omicron infection beyond that afforded by primary vaccination among people with and without a prior documented infection.

## Methods

### Study Setting and Population

We conducted a test-negative case-control (TNCC) analysis using data collected as part of the Studying COVID-19 Outcomes after SARS-CoV-2 Infection and Vaccination (SUCCESS) Study at the Yale New Haven Health System (YNHH). The YNHH is a large academic health system comprising five hospital delivery networks and associated outpatient clinics in Connecticut and Rhode Island. We chose the TNCC design because it has been shown to provide effectiveness estimates consistent with those from randomized control trials, has been widely applied to estimate real-world effectiveness for COVID-19 vaccines, and mitigates the risk of confounding introduced by care-seeking and testing access.(1,12–15)

The study population comprised vaccine eligible (≥5 years of age) people who had at least one SARS-CoV-2 test or mRNA (mRNA-1273 [Moderna] or BNT162b2 [Pfizer]) vaccine dose in the medical records. We identified SARS-CoV-2 RT-PCR tests that were collected from the study population and performed with the TaqPath™ COVID⍰19 (Thermo Fisher Scientific) diagnostic assay between November 1, 2021 and January 31, 2022, the period prior to and during the Omicron epidemic wave in Connecticut (Figure 1A). At the beginning of the study, Delta was the predominant variant in Connecticut, accounting for 99.63% (3,808 of 3,822) of the sequenced samples deposited in the GISAID database that were collected between November 1 and November 28, 2021.(16) We used the TaqPath assay to select tests as cases and controls since its S-gene probe, which fails for Omicron (BA.1) but not for Delta, allows for prediction of an Omicron infection when the primary circulating variants are Omicron and Delta.(17)

**Figure 1:**
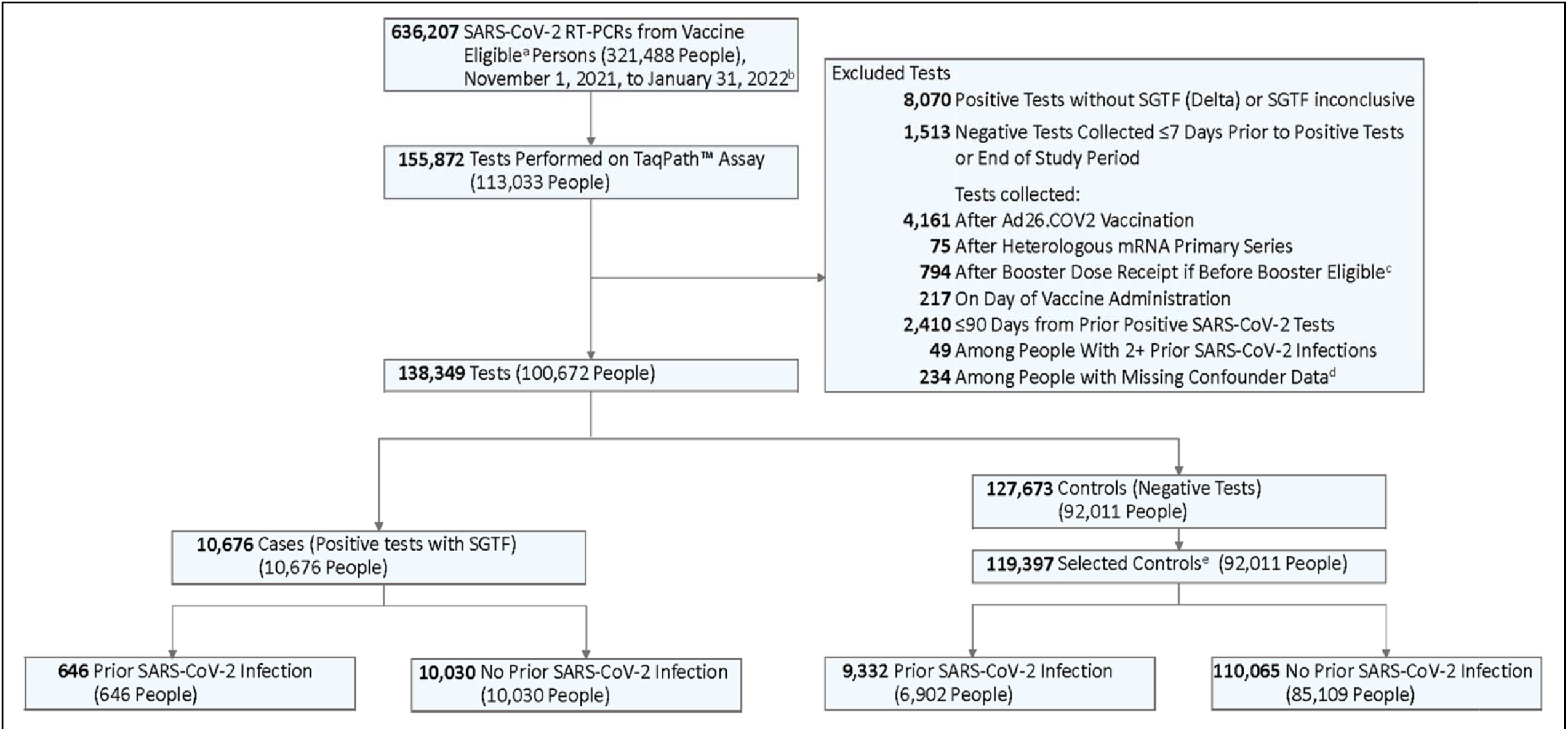
Selection of Tests for the Case Control Analysis. Legend: The sample was limited to RT-PCRs run on the TaqPath™ COVID⍰19 (Thermo Fisher Scientific) among vaccine eligible individuals. Case status was defined based on the reflex results. We included all positive tests (cases) and up to three negative tests (controls) per person. Cases and controls were stratified by presence of prior infection, or a positive RT-PCR or rapid antigen test at least 90 days before testing. ^a^ Vaccine eligible defined as age ≥5 years; ^b^ The first S-gene target failure (SGTF) defined Omicron (BA.1 Lineage) variant infection in the study population was identified on November 11, 2021; ^c^ Excluded tests that were performed after a person was given a booster dose before FDA authorization or that was given less than 150 days after primary vaccination completion; ^d^ There were 182 people with missing SVI data and 52 people with missing sex data; ^e^ People were allowed to contribute up to three negative tests to the control sample. If they had more than three negative tests over the study period, three tests were randomly selected. If a person had more than one negative test within a 7-day period, a random test was selected.

We excluded tests that were performed after receiving a heterologous primary vaccination (i.e. different first and second dose manufacturers) or an Ad26.COV2 vaccine dose. Additionally, we excluded tests that were performed among people who received booster doses prior to eligibility (defined as five months since 2^nd^ primary vaccine dose and after booster vaccination approval in the US [September 22, 2021]).^16^ We excluded tests that were performed in the 90 days after a positive SARS-CoV-2 test (rapid antigen or RT-PCR), had a positive reflex result with an inconclusive S-gene target failure (SGTF) finding, or were obtained from people with more than one prior SARS-CoV-2 infection or with missing confounder data (Figure 1).

The Yale Computational Health Platform was used to extract demographic, comorbidity,

COVID-19 vaccination and SARS-CoV-2 testing data from electronic medical records.(18,19) Additional COVID-19 vaccination records from the state vaccination registry were linked to the YNHH medical records and extracted through the same platform. This study was approved by the Yale Institutional Review Board (ID# 2000030222).

### Exposures

Our exposure of interest was time from completion of primary (two doses) and booster (third dose) vaccination with mRNA-1273 or BNT162b2. We stratified vaccination by time since dose (<14 days since 2^nd^ dose, 14-149 days since 2^nd^ dose, ≥150 days since 2^nd^ dose but prior to booster dose, <14 days since booster dose, 14-59 days since booster dose, 60-89 days since booster dose and ≥90 days since booster dose). Tests were further stratified by a history of a prior SARS-CoV-2 infection, defined as a positive RT-PCR or rapid antigen test result in the medical record ≥90 days before the included test.

### Case and Control Definition and Selection

A case of Omicron infection was a positive SARS-CoV-2 RT-PCR test with SGTF, defined as: 1] an ORF1ab Ct value of < 30 and S-gene Ct – ORF1ab Ct value ≥5; or 2] ORF1ab Ct value <30 and S-gene Ct value ≥40.(17,20) An eligible control was a negative SARS-CoV-2 RT-PCR test collected ≥7 days prior to a positive test or the end of the study period (to account for non-resulted cases). Our sample included all eligible cases and up to three negative tests (controls) per person during the study period. If an individual had more than one negative test within a seven-day period, one random test was selected during the period as a control.

### Statistical Analysis

We conducted two primary analyses, each stratified by prior SARS-CoV-2 infection status. First, we estimated vaccine effectiveness as one minus the odds ratio for infection comparing vaccinated to unvaccinated people. Second, we examined whether a booster dose was associated with increased protection beyond that afforded by the primary series by comparing the odds of infection among recently boosted people (14-59 days after booster dose) to the odds among booster eligible people. In alignment with CDC booster dose recommendations at the time of analysis,(21) we defined booster eligible people as those who completed their primary series ≥150 days (five months) prior to the included test and had not received a booster dose. For this analysis, we were interested in the level of protection associated with a booster dose. For that reason, we excluded tests that were collected among boosted people who experienced a previous SARS-CoV-2 infection after receiving their booster dose.

As a secondary analysis, we evaluated whether the odds of infection changed over time after the administration of a booster dose by comparing the odds of infection among recently boosted people (14-59 days after booster dose) to the odds of infection among people who received their booster dose 60-89 and ≥90 days prior to testing.(22) Further, to test if changes in the odds of infection over time since booster dose receipt resulted in a loss of protection relative to booster eligible individuals, we compared the odds of infection among people who received their booster dose 60-89 and ≥90 days prior to testing to the odds of infection among booster eligible people. Since there was a limited number of boosted people with a prior SARS-CoV-2 infection (n=651), the secondary analysis was restricted to people without a prior infection.

A mixed effects generalized additive logistic regression was used to evaluate associations. We included the following *a priori* selected covariates: date of test (continuous), age (continuous), sex, race/ethnicity, Charlson comorbidity score (categorized as 0, 1-2, 3-4, 5+)(23), number of non-emergent YNHH encounters in the year prior to vaccine rollout in Connecticut (December 2020; categorized as 0, 1-2, 3-4, 5+), insurance group (uninsured, Medicaid, Medicare, other), social vulnerability index (SVI) of residential zip code (continuous)^22^ and municipality. Continuous factors were modeled using a natural spline with 3 knots and we included a random intercept for municipality.(24,25) To account for waning infection-mediated immunity, we included time since prior SARS-CoV-2 infection as a continuous factor in analyses limited to people with prior infection. All analyses were conducted in R, version 4.1.2.(26)

### Sensitivity Analyses

We performed multiple sensitivity analyses to ensure our findings were robust to alternative study design, data cleaning and modeling assumptions. Specifically, we tested the robustness of our findings to the following scenarios: 1:1 matching with replacement, exclusion of heterologous booster doses, inclusion of tests among people with more than one prior infection, exclusion of discordant test results, inclusion of positive TaqPath results with inconclusive SGTF (included as negative tests), and inclusion of all controls. To examine if the temporality of prior SARS-CoV-2 infection and vaccinations impacted estimates of vaccine effectiveness among people with a prior infection, we conducted an analysis where we excluded tests performed among people whose prior SARS-CoV-2 infection occurred after the first dose of primary vaccination. Additionally, to evaluate whether waning of protection associated with primary vaccination influenced the risk comparisons between boosted and booster eligible people, we conducted sensitivity analyses that were restricted to tests collected among people who completed primary vaccination ≥150 days prior to testing and adjusted for time since completion of primary vaccination. For a detailed description, see the section on *Sensitivity Analyses* in the *Supplement*.

## Results

### Study Population

Between November 1, 2021, and January 31, 2022, we identified 155,827 SARS-CoV-2 tests that were performed with the TaqPath assay on samples obtained from 113,033 unique people in the YNHH system (Figure 1). The first SGTF defined Omicron infection in the study population was identified on November 11, 2021 (Figure 2). Of the 138,349 eligible tests, 10,676 were identified as Omicron (BA.1) infections (cases). From the 127,673 negative RT-PCRs, we randomly selected up to three negative tests (controls) per person, resulting in 119,397 controls (Figure 1).

**Figure 2:**
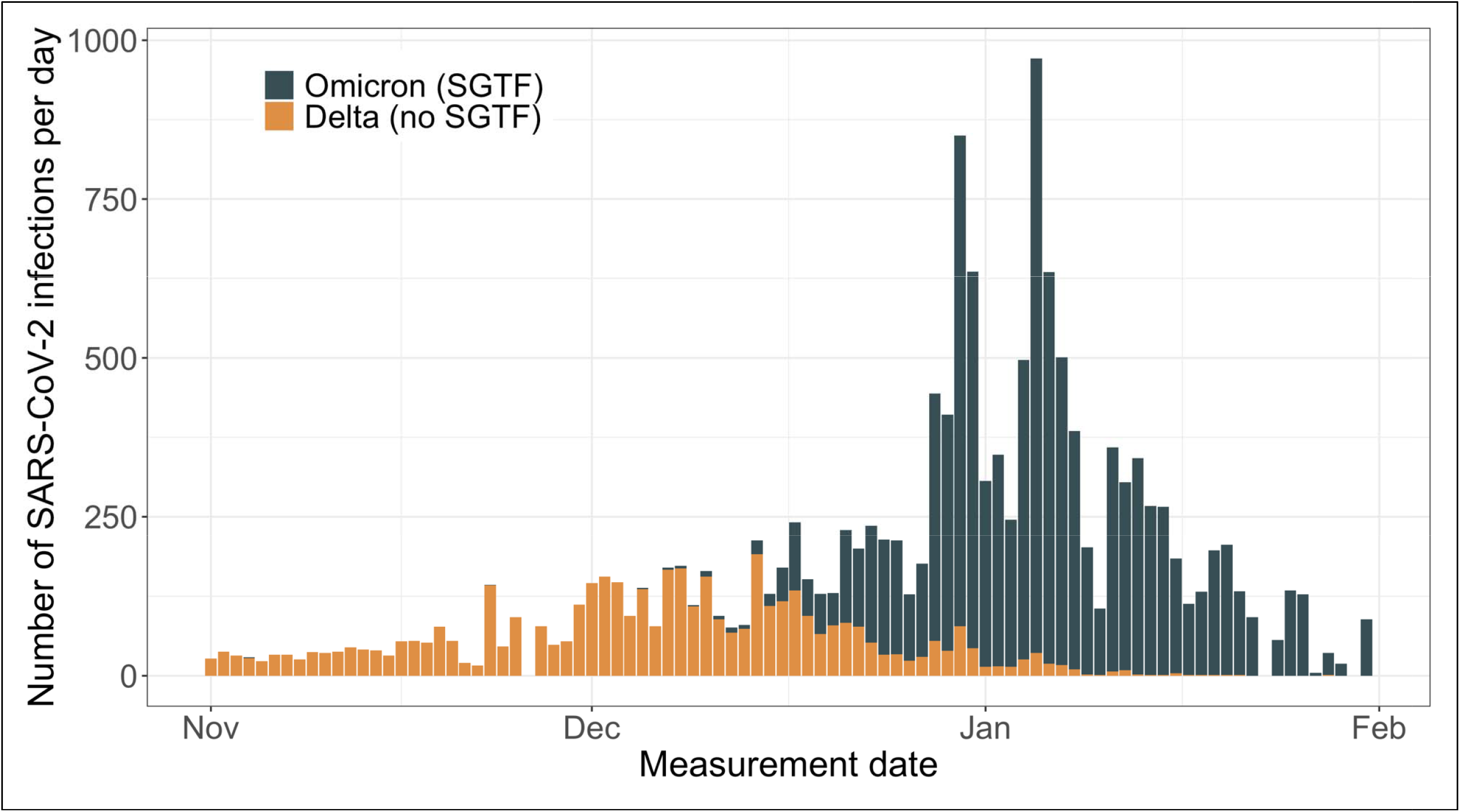
Daily SARS-CoV-2 Infections Caused by Omicron (BA. 1 Lineage) and Delta Variants Identified during TaqPath Testing at the Yale New Haven Hospital System between November 1, 2021 and January 31, 2022. Legend: TaqPath™ COVID⍰19 (Thermo Fisher Scientific) confirmed SARS-CoV-2 infections among vaccine eligible individuals. Infections were classified as Omicron (BA.1 Lineage) based on presence of S-gene target failure (SGTF).

Cases and controls had similar characteristics with respect to age, gender, SVI of residential zip code and Charlson comorbidity score (Table 1). However, a larger proportion of Omicron cases occurred among non-Hispanic Black people (16.6% vs 10.1% in controls) and those who were uninsured (10.2% vs 7.6% in controls) or received Medicaid (23.7% vs 13.4% in controls). Among boosted people, the median time between booster vaccination and testing was similar for cases (53 days [1^st^-3^rd^ Quartile: 27 to 77 days]) and controls (41 days [1^st^-3^rd^ Quartile: 22 to 64 days]). Among cases and controls, 6.1% and 7.8% had a prior SARS-CoV-2 infection. The time between prior infection and testing was similar for cases (372 days [1^st^-3^rd^ Quartile: 295 to 418 days]) and controls (328 days [1^st^-3^rd^ Quartile: 258 to 384 days]).

**Table 1:**
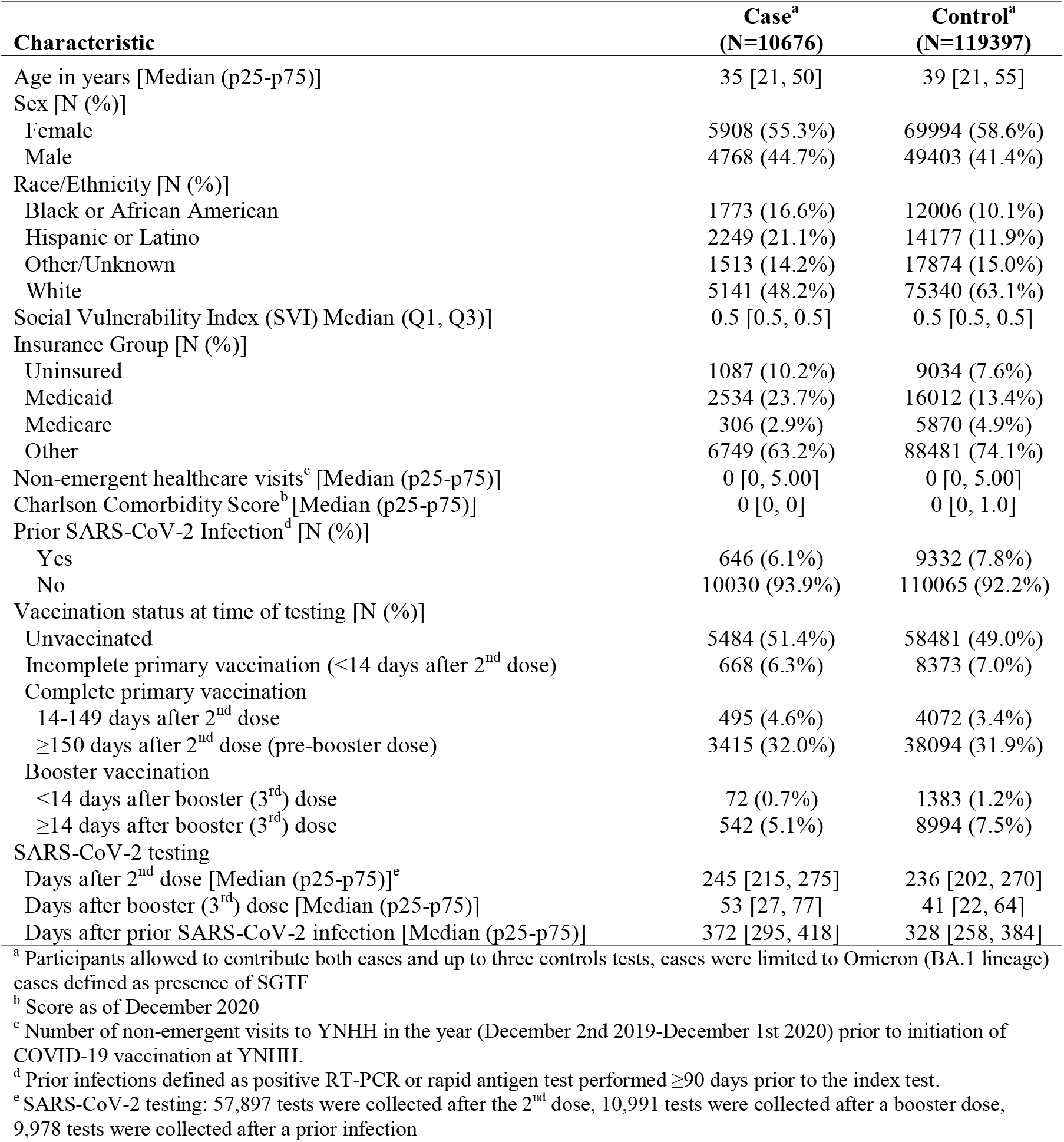
Characteristics of SARS-CoV-2 Tests Included as Cases or Controls between November 1, 2021 and January 31, 2022.

| Characteristic | Case<sup>a</sup><br>(N=10676) | Control<sup>a</sup><br>(N=119397) |
| --- | --- | --- |
| Age in years [Median (p25-p75)] | 35 [21, 50] | 39 [21, 55] |
| Sex [N (%)] |  |  |
| Female | 5908 (55.3%) | 69994 (58.6%) |
| Male | 4768 (44.7%) | 49403 (41.4%) |
| Race/Ethnicity [N (%)] |  |  |
| Black or African American | 1773 (16.6%) | 12006 (10.1%) |
| Hispanic or Latino | 2249 (21.1%) | 14177 (11.9%) |
| Other/Unknown | 1513 (14.2%) | 17874 (15.0%) |
| White | 5141 (48.2%) | 75340 (63.1%) |
| Social Vulnerability Index (SVI) Median (Q1, Q3)] | 0.5 [0.5, 0.5] | 0.5 [0.5, 0.5] |
| Insurance Group [N (%)] |  |  |
| Uninsured | 1087 (10.2%) | 9034 (7.6%) |
| Medicaid | 2534 (23.7%) | 16012 (13.4%) |
| Medicare | 306 (2.9%) | 5870 (4.9%) |
| Other | 6749 (63.2%) | 88481 (74.1%) |
| Non-emergent healthcare visits<sup>c</sup> [Median (p25-p75)] | 0 [0, 5.00] | 0 [0, 5.00] |
| Charlson Comorbidity Score<sup>b</sup> [Median (p25-p75)] | 0 [0, 0] | 0 [0, 1.0] |
| Prior SARS-CoV-2 Infection<sup>d</sup> [N (%)] |  |  |
| Yes | 646 (6.1%) | 9332 (7.8%) |
| No | 10030 (93.9%) | 110065 (92.2%) |
| Vaccination status at time of testing [N (%)] |  |  |
| Unvaccinated | 5484 (51.4%) | 58481 (49.0%) |
| Incomplete primary vaccination (<14 days after 2<sup>nd</sup> dose) | 668 (6.3%) | 8373 (7.0%) |
| Complete primary vaccination |  |  |
| 14-149 days after 2<sup>nd</sup> dose | 495 (4.6%) | 4072 (3.4%) |
| ≥150 days after 2<sup>nd</sup> dose (pre-booster dose) | 3415 (32.0%) | 38094 (31.9%) |
| Booster vaccination |  |  |
| <14 days after booster (3<sup>rd</sup>) dose | 72 (0.7%) | 1383 (1.2%) |
| ≥14 days after booster (3<sup>rd</sup>) dose | 542 (5.1%) | 8994 (7.5%) |
| SARS-CoV-2 testing |  |  |
| Days after 2<sup>nd</sup> dose [Median (p25-p75)]<sup>e</sup> | 245 [215, 275] | 236 [202, 270] |
| Days after booster (3<sup>rd</sup>) dose [Median (p25-p75)] | 53 [27, 77] | 41 [22, 64] |
| Days after prior SARS-CoV-2 infection [Median (p25-p75)] | 372 [295, 418] | 328 [258, 384] |

<sup>a</sup> Participants allowed to contribute both cases and up to three controls tests, cases were limited to Omicron (BA.1 lineage) cases defined as presence of SGTF
<sup>b</sup> Score as of December 2020
<sup>c</sup> Number of non-emergent visits to YNHH in the year (December 2nd 2019-December 1st 2020) prior to initiation of COVID-19 vaccination at YNHH.
<sup>d</sup> Prior infections defined as positive RT-PCR or rapid antigen test performed ≥90 days prior to the index test.
<sup>e</sup> SARS-CoV-2 testing: 57,897 tests were collected after the 2<sup>nd</sup> dose, 10,991 tests were collected after a booster dose, 9,978 tests were collected after a prior infection

### Risk of Omicron Infection among Boosted and Booster Eligible People

During the period prior to booster eligibility (14-149 days after 2^nd^ dose), the effectiveness of primary mRNA vaccination against Omicron infection was 36.1% (95% Confidence Intervals [CI], 7.1% to 56.1%) and 28.5% (95% CI, 20.0% to 36.2%) for people with and without a prior infection, respectively. During the period of booster eligibility (150+ days after 2^nd^ dose), the effectiveness of primary vaccination was 34.0% (95% CI, 18.5% to 46.5%) for people with and 15.3% (95% CI, 10.4% to 20.2%) for people without a prior infection. Vaccine effectiveness in the period 14-59 days after a booster dose was 45.8% (95% CI, 20.0% to 63.2%) and 56.9% (95% CI, 52.1% to 61.2%) for people with and without a prior infection, respectively (Figure 3).

**Figure 3:**
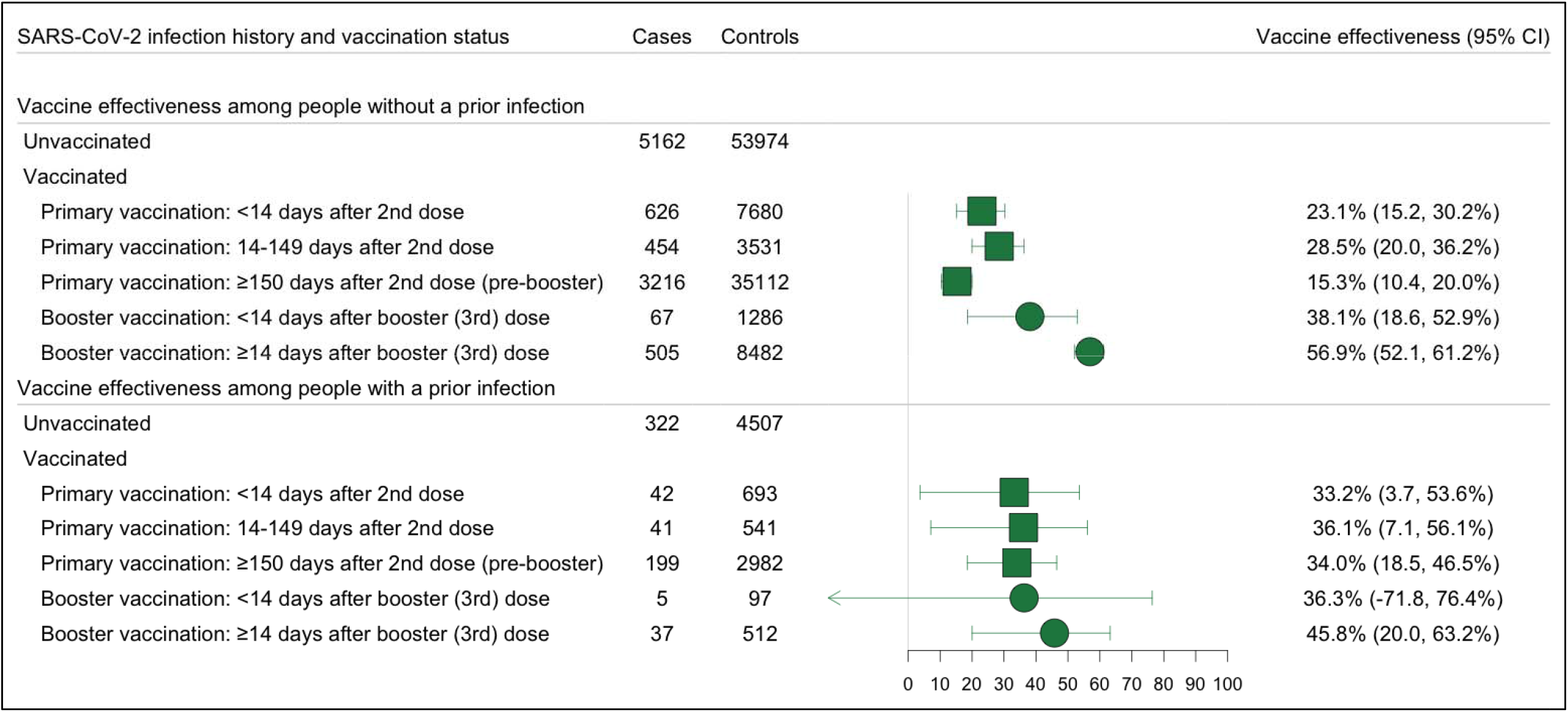
Effectiveness of Primary and Booster Vaccination with COVID-19 mRNA Vaccines Against SARS-COV-2 Omicron (BA. 1 Lineage) Variant Infections, Stratified by the History of a Prior SARS-CoV-2 Infection. Legend: Forest plot depicting vaccine effectiveness against any Omicron (BA.1 Lineage) and Delta infections for both US approved mRNA vaccines (BNT162b2 and mRNA-1273) among people with and without a prior infection. Prior infection was defined as a positive RT-PCR or rapid antigen test at least 90 days before testing. Omicron infection was defined as the presence of S-gene target failure (SGTF) defined as ORF1ab Ct < 30 and S-gene – ORF1ab >= 5, or ORF1ab < 30 and S-gene >= 40. Vaccine effectiveness was estimated as 1-OR from a model adjusted for date of test, age, sex, race/ethnicity, Charlson comorbidity score, number of non-emergent visits in the year prior to the vaccine rollout in Connecticut, insurance status, municipality, and social venerability index (SVI) of residential zip code in all analyses and time between testing and last prior infection in analyses of people with prior infection.

The odds of Omicron infection did not differ significantly between boosted and booster eligible people with a prior SARS-CoV-2 infection (OR: 0.83 [95% CI, 0.56 to 1.22]). However, among people without a prior infection, the odds of infection were 0.51 (95% CI, 0.46 to 0.56) times lower for boosted than booster eligible people (Table 2).

**Table 2:**
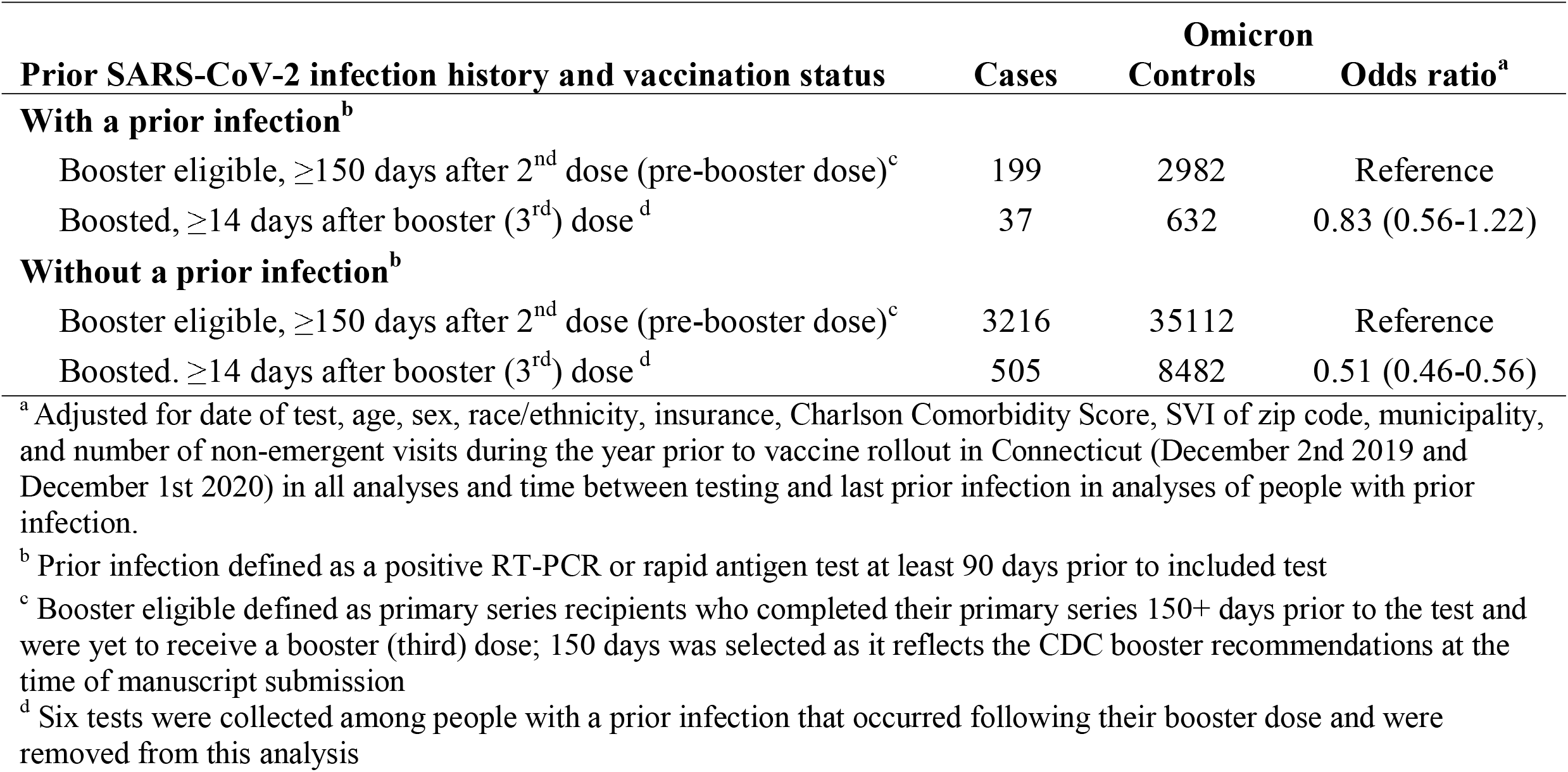
Risk of SARS-CoV-2 Omicron (BA.1 Lineage) Variant Infection among People Who Received Booster Vaccination Relative to Booster Eligible People, According to History of a Prior SARS-CoV-2 Infection

In the secondary analysis that was restricted to people without a prior infection, the odds of Omicron infection increased over time since booster vaccination and was significantly higher 90+ days after a booster dose relative to the period 14-59 days after the dose (OR: 1.6 [95% CI, 1.2 to 2.0], Supplemental Table 1). Yet, the odds of infection among boosted people 90+ days after the booster dose was lower than the odds among booster eligible people (OR: 0.72 [95% CI, 0.66 to 0.91], Table 3).

**Table 3.**
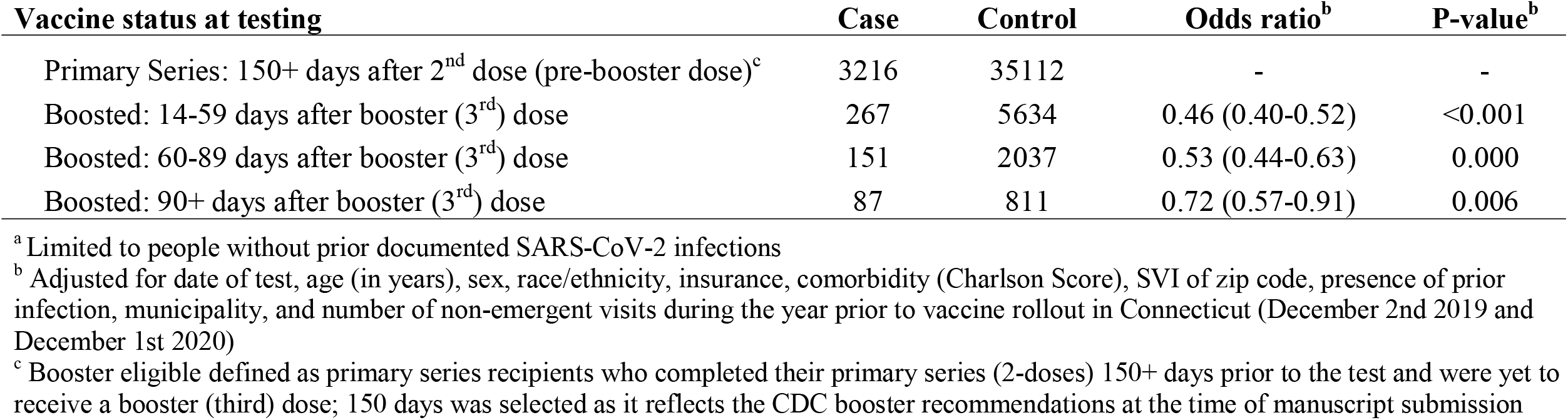
Risk of SARS-CoV-2 Omicron (BA.1 Lineage) Variant Infection among People Who Received Booster Vaccination Relative to Booster Eligible People^a^, According to Time after Receiving a Booster Vaccine Dose

| **Vaccine status at testing** | **Case** | **Control** | **Odds ratio<sup>b</sup>** | **P-value<sup>b</sup>** |
| --- | --- | --- | --- | --- |
| Primary Series: 150+ days after 2<sup>nd</sup> dose (pre-booster dose)<sup>c</sup> | 3216 | 35112 | - | - |
| Boosted: 14-59 days after booster (3<sup>rd</sup>) dose | 267 | 5634 | 0.46 (0.40-0.52) | <0.001 |
| Boosted: 60-89 days after booster (3<sup>rd</sup>) dose | 151 | 2037 | 0.53 (0.44-0.63) | 0.000 |
| Boosted: 90+ days after booster (3<sup>rd</sup>) dose | 87 | 811 | 0.72 (0.57-0.91) | 0.006 |

<sup>a</sup> Limited to people without prior documented SARS-CoV-2 infections
<sup>b</sup> Adjusted for date of test, age (in years), sex, race/ethnicity, insurance, comorbidity (Charlson Score), SVI of zip code, presence of prior infection, municipality, and number of non-emergent visits during the year prior to vaccine rollout in Connecticut (December 2nd 2019 and December 1st 2020)
<sup>c</sup> Booster eligible defined as primary series recipients who completed their primary series (2-doses) 150+ days prior to the test and were yet to receive a booster (third) dose; 150 days was selected as it reflects the CDC booster recommendations at the time of manuscript submission

### Sensitivity Analyses

In sensitivity analyses, the effectiveness of booster vaccination against Omicron infection (≥14 days after the booster dose) ranged between 35.1% and 48.5% for people with a prior infection and 55.0% and 58.5% for people without a prior infection (Supplemental Figures 1-8). Compared with the primary analysis, we observed lower precision from our matched (1:1 with replacement) analysis (Supplemental Figure 1). Following the exclusion of tests collected among people whose prior infection occurred after their first vaccine dose, we observed non-significantly lower effectiveness estimates for primary (14-149 days after 2^nd^ dose: 33.5% [95% CI, 2.7% to 54.6%]) and booster (14+days after booster dose: 41.9% [95% CI, 12.1% to 61.6%]) vaccination. Adjusting for time between testing and completion of primary vaccination did not significantly alter the estimated association between booster doses and Omicron infections among people with a prior infection (OR: 0.81 [95% CI, 0.53 to 1.23]) or without prior infection (OR: 0.55 [CI, 0.49 to 0.61]) (Supplemental Table 2).

## Discussion

Leveraging data from a large population of Connecticut residents, we examined the effectiveness of primary series and booster doses against Omicron infections among primary series (2-dose) mRNA recipients with and without a prior infection. We found primary vaccination was associated with significant but low levels of protection among people with and without a prior infection. While booster vaccination was associated with protection beyond that afforded by the primary series in people without a prior infection, we did not identify a significant increase in protection among people with a prior infection.

Contrary to the findings of Shrestha et al.,(11) our analysis, which ascertained Omicron infection in cases by the presence of SGTF and had increased precision for vaccine effectiveness, found that primary vaccination was associated with a significant reduction in the risk (34.0% [95% CI, 18.5% to 46.5%]) of Omicron infection among people with a prior infection. Though we found the level of protection afforded by primary vaccination to be low, our findings suggest that primary vaccination may be warranted regardless of prior infection status.

Our estimate of booster vaccination effectiveness among previously infected people had reduced precision (45.8% [95% CI, 20.0% to 63.2%]) compared to the estimate for people without a prior infection (56.9% [95% CI, 52.1% to 61.2%]). Because the differences in these estimates may be driven by differences in care-seeking behaviors, we caution their direct comparison. However, in a parallel analysis, the odds of infection did not differ significantly (OR: 0.83 [95% CI, 0.56 to 1.22]) between boosted and booster eligible people with a prior SARS-CoV-2 infection. Together, these findings suggest that boosters may not confer additional protection beyond that afforded by primary vaccination among previously infected people and that, in vaccine limited scenarios, individuals who received a primary series and have been previously infected should not be prioritized for a booster dose. These findings provide supportive evidence for the inclusion of documented prior SARS-CoV-2 infections in addition to vaccinations for future vaccination requirements and mandatory proof of immunity (such as vaccination cards). (11,27–32) However, given the reduced precision of these estimates, additional research from other regions should be conducted to provide additional clarity on the benefits of boosters within this sub-population.

In alignment with prior studies,(1,7,33) we found the risk of Omicron infection among boosted people without a prior infection increased significantly three months after booster dose administration. However, the odds of infection among boosted people remained significantly lower than the odds among booster eligible people (OR: 0.7 [95% CI, 0.6 to 0.9]). Thus, even with the decline in protection, booster vaccination appears to provide additional protection beyond that conferred by primary vaccination in people without a prior SARS-CoV-2 infection.

Weekly testing for certain unvaccinated professionals, such as employees of healthcare facilities that accept Medicare and/or Medicaid or Connecticut state employees, was required by the state and federal government during the study period.(34,35) Because such requirements resulted in increased testing among unvaccinated but not vaccinated persons, our vaccine effectiveness estimates are likely conservative. However, the bias introduced by required testing does not extend to the comparisons among vaccinated groups and our findings comparing boosted to booster eligible people are likely to be unaffected by testing policies.

## Limitations

Our analysis was limited to a population of Connecticut residents and was reliant on medical record data that is subject to misclassification. In the place of whole genome sequence data, we used SGTF status to ascertain Omicron infections as cases. The use of SGTF as a proxy has been widely used during the Omicron epidemic wave and has been recommended as an indicator of Omicron lineage BA.1 infection by the WHO.^1,19^ While a new sub-lineage of the Omicron variant (BA.2) has been detected in the US without SGTF, sequencing data from YNHH showed 100% agreement between SGTF and whole genome sequence-defined Omicron through December 2021.(17) In January 2022, we observed a small number (184) of positive tests that did not have SGTF and were not included as cases in the analyses. Our sample is overly representative of mild cases of SARS-CoV-2 infection since TaqPath testing was primarily employed in the YNHH outpatient setting. Additionally, our sample excluded cases with high Ct values since Ct values of at least 30 were required for SGTF calls.

We did not have adequate sample to evaluate the level of protection conferred by two or more prior infections (n=49). We expect a proportion of prior SARS-CoV-2 infections may have gone undetected and that ascertainment of prior infection history may be subject to misclassification.

Nevertheless, our analyses of people with documented prior infection are unaffected by such misclassification. Despite accounting for health seeking behavior in our study design and confounder selection, residual behavioral differences may exist. Finally, the analyses were not powered to test associations for severe COVID-19, we therefore cannot exclude that booster vaccination may increase protection against such outcomes in people with a prior SARS-CoV-2 infection.

## Conclusion

Primary vaccination with two COVID-19 mRNA vaccine doses provided significant but limited protection among people with and without a prior SARS-CoV-2 infection. While booster vaccination resulted in additional protection beyond that afforded by the primary vaccination among people without a prior infection, it did not result in additional protection among previously infected people. These findings support primary vaccination regardless of prior infection history but suggest that a person’s history of prior SARS-CoV-2 infection should be considered in subsequent vaccination decisions, such as booster vaccination.

## Data Availability

The data used in this study belongs to Yale University. Qualified researchers may submit a data share request for de-identified patient level data by contacting the corresponding author with a detailed description of the research question.

## Funding

Funding for the Studying COVID-19 Outcomes after SARS-CoV-2 Infection and Vaccination (SUCCESS) Study was provided by the Beatrice Kleinberg Neuwirth and Sendas Family Funds, Merck and Co through their Merck Investigator Studies Program, and the Yale Schools of Public Health and Medicine.

## Author Contributions

Margaret L. Lind, Albert I. Ko and Wade L. Schulz have full access to all the data in the study and take responsibility for the integrity of the data and the accuracy of the data analysis. *Concept and design:* Lind, Hitchings, Robertson, Schulz, Ko, Cummings, Silva

*Acquisition and interpretation of data:* Lind, Warner, Coppi, Robertson, Duckwell

*Drafting of the manuscript:* Lind, Robertson, Hitchings, Ko

*Critical revision of the results:* All authors.

*Critical revision of the manuscript for important intellectual content:* All authors.

*Statistical analysis:* Lind, Robertson

*Administrative, technical, or material support:* Duckwall, Borg, Giuseppe, Sosensky

*Supervision:* Ko, Hitchings, Schulz, Cummings, Iwasaki

## Conflict of Interest Disclosures

A.I.K serves as an expert panel member for Reckitt Global Hygiene Institute, scientific advisory board member for Revelar Biotherapeutics and a consultant for Tata Medical and Diagnostics and Regeneron Pharmaceuticals, and has received grants from Merck, Regeneron Pharmaceuticals and Tata Medical and Diagnostics for research related to COVID-19. W.L.S. was an investigator for a research agreement, through Yale University, from the Shenzhen Center for Health Information for work to advance intelligent disease prevention and health promotion; collaborates with the National Center for Cardiovascular Diseases in Beijing; is a technical consultant to Hugo Health, a personal health information platform, and co-founder of Refactor Health, an AI-augmented data management platform for healthcare; and has received grants from Merck and Regeneron Pharmaceutical for research related to COVID-19. Other authors declare no conflict of interest.

## Notes

### Author Declarations

This study was approved by the Yale Institutional Review Board (ID# 2000030222).

### Summary of Updates

Updated abstract and figure/table formatting.

## References

1. Accorsi EK, Britton A, Fleming-Dutra KE, Smith ZR, Shang N, Derado G, et al. Association Between 3 Doses of mRNA COVID-19 Vaccine and Symptomatic Infection Caused by the SARS-CoV-2 Omicron and Delta Variants. JAMA [Internet]. 2022 Jan 21 [cited 2022 Jan 26]; Available from: https://doi.org/10.1001/jama.2022.0470

2. Andrews N, Stowe J, Kirsebom F, Toffa S, Rickeard T, Gallagher E, et al. Effectiveness of COVID-19 vaccines against the Omicron (B.1.1.529) variant of concern [Internet]. medRxiv; 2021 [cited 2022 Jan 26]. p. 2021.12.14.21267615. Available from: https://www.medrxiv.org/content/10.1101/2021.12.14.21267615v1

3. Johnson AG. COVID-19 Incidence and Death Rates Among Unvaccinated and Fully Vaccinated Adults with and Without Booster Doses During Periods of Delta and Omicron Variant Emergence — 25 U.S. Jurisdictions, April 4–December 25, 2021. MMWR Morb Mortal Wkly Rep [Internet]. 2022 [cited 2022 Jan 26];71. Available from: https://www.cdc.gov/mmwr/volumes/71/wr/mm7104e2.htm

4. Thompson MG. Effectiveness of a Third Dose of mRNA Vaccines Against COVID-19–Associated Emergency Department and Urgent Care Encounters and Hospitalizations Among Adults During Periods of Delta and Omicron Variant Predominance — VISION Network, 10 States, August 2021–January 2022. MMWR Morb Mortal Wkly Rep [Internet]. 2022 [cited 2022 Jan 26];71. Available from: https://www.cdc.gov/mmwr/volumes/71/wr/mm7104e3.htm

5. Sheikh A, McMenamin J, Taylor B, Robertson C. SARS-CoV-2 Delta VOC in Scotland: demographics, risk of hospital admission, and vaccine effectiveness. The Lancet. 2021 Jun 26;397(10293):2461–2.

6. Hall V, Foulkes S, Insalata F, Saei A, Kirwan P, Atti A, et al. Effectiveness and durability of protection against future SARS-CoV-2 infection conferred by COVID-19 vaccination and previous infection; findings from the UK SIREN prospective cohort study of healthcare workers March 2020 to September 2021 [Internet]. medRxiv; 2021 [cited 2022 Jan 26]. p. 2021.11.29.21267006. Available from: https://www.medrxiv.org/content/10.1101/2021.11.29.21267006v1

7. Hall V, Foulkes S, Insalata F, Kirwan P, Saei A, Atti A, et al. Protection against SARS-CoV-2 after Covid-19 Vaccination and Previous Infection. N Engl J Med. 2022 Feb 16;0(0):null.

8. Nordström P, Ballin M, Nordström A. Risk of SARS-CoV-2 reinfection and COVID-19 hospitalisation in individuals with natural and hybrid immunity: a retrospective, total population cohort study in Sweden. Lancet Infect Dis [Internet]. 2022 Mar 31 [cited 2022 Mar 31];0(0). Available from: https://www.thelancet.com/journals/laninf/article/PIIS1473-3099(22)00143-8/fulltext

9. León TM. COVID-19 Cases and Hospitalizations by COVID-19 Vaccination Status and Previous COVID-19 Diagnosis — California and New York, May–November 2021. MMWR Morb Mortal Wkly Rep [Internet]. 2022 [cited 2022 Feb 14];71. Available from: https://www.cdc.gov/mmwr/volumes/71/wr/mm7104e1.htm

10. Cerqueira-Silva T, Andrews JR, Boaventura VS, Ranzani OT, Oliveira V de A, Paixão ES, et al. Effectiveness of CoronaVac, ChAdOx1, BNT162b2 and Ad26.COV2.S among individuals with prior SARS-CoV-2 infection in Brazil [Internet]. medRxiv; 2021 [cited 2022 Jan 26]. p. 2021.12.21.21268058. Available from: https://www.medrxiv.org/content/10.1101/2021.12.21.21268058v1

11. Shrestha NK, Burke PC, Nowacki AS, Terpeluk P, Gordon SM. Necessity of COVID-19 Vaccination in Persons Who Have Already Had COVID-19. Clin Infect Dis Off Publ Infect Dis Soc Am. 2022 Jan 13;ciac022.

12. De Serres G, Skowronski DM, Wu XW, Ambrose CS. The test-negative design: validity, accuracy and precision of vaccine efficacy estimates compared to the gold standard of randomised placebo-controlled clinical trials. Euro Surveill Bull Eur Sur Mal Transm Eur Commun Dis Bull. 2013 Sep 12;18(37):20585.

13. Dean NE, Longini I. Lecture 9: Study designs for evaluating vaccine efficacy [Internet]. University of Florida. Available from: https://si.biostat.washington.edu/sites/default/files/modules/2019_SISMID_06_9.pdf

14. Hitchings MDT, Ranzani OT, Dorion M, D’Agostini TL, de Paula RC, de Paula OFP, et al. Effectiveness of ChAdOx1 vaccine in older adults during SARS-CoV-2 Gamma variant circulation in São Paulo. Nat Commun. 2021 Oct 28;12(1):6220.

15. Ranzani OT, Hitchings MDT, Dorion M, D’Agostini TL, de Paula RC, de Paula OFP, et al. Effectiveness of the CoronaVac vaccine in older adults during a gamma variant associated epidemic of covid-19 in Brazil: test negative case-control study. The BMJ. 2021 Aug 20;374:n2015.

16. Elbe S, Buckland-Merrett G. Data, disease and diplomacy: GISAID’s innovative contribution to global health. Glob Chall. 2017;1(1):33–46.

17. Chaguza C, Coppi A, Earnest R, Ferguson D, Kerantzas N, Warner F, et al. Rapid emergence of SARS-CoV-2 Omicron variant is associated with an infection advantage over Delta in vaccinated persons [Internet]. medRxiv; 2022 [cited 2022 Mar 11]. p. 2022.01.22.22269660. Available from: https://www.medrxiv.org/content/10.1101/2022.01.22.22269660v1

18. McPadden J, Durant TJ, Bunch DR, Coppi A, Price N, Rodgerson K, et al. Health Care and Precision Medicine Research: Analysis of a Scalable Data Science Platform. J Med Internet Res. 2019 Apr 9;21(4):e13043.

19. Schulz WL, Durant TJS, Jr CJT Hsiao AL, Krumholz HM. Agile Health Care Analytics: Enabling Real-Time Disease Surveillance With a Computational Health Platform. J Med Internet Res. 2020 May 28;22(5):e18707.

20. European Centre for Disease Prevention and Control, Europe WHORO for. Methods for the detection and identification of SARS-CoV-2 variants, March 2021 [Internet]. World Health Organization. Regional Office for Europe; 2021 [cited 2022 Apr 19]. Report No.: WHO/EURO:2021-2148-41903-57493. Available from: https://apps.who.int/iris/handle/10665/340067

21. CDC. COVID-19 Booster Shot [Internet]. Centers for Disease Control and Prevention. 2022 [cited 2022 Apr 5]. Available from: https://www.cdc.gov/coronavirus/2019-ncov/vaccines/booster-shot.html

22. Lipsitch M, Goldstein E, Ray GT, Fireman B. Depletion-of-susceptibles bias in influenza vaccine waning studies: how to ensure robust results. Epidemiol Infect. 2019 Nov 27;147:e306.

23. Charlson ME, Pompei P, Ales KL, MacKenzie CR. A new method of classifying prognostic comorbidity in longitudinal studies: development and validation. J Chronic Dis. 1987;40(5):373–83.

24. Kahan BC, Rushton H, Morris TP, Daniel RM. A comparison of methods to adjust for continuous covariates in the analysis of randomised trials. BMC Med Res Methodol. 2016 Apr 11;16(1):42.

25. Groenwold RHH, Klungel OH, Altman DG, van der Graaf Y, Hoes AW, Moons KGM. Adjustment for continuous confounders: an example of how to prevent residual confounding. CMAJ Can Med Assoc J. 2013 Mar 19;185(5):401–6.

26. R Core Team. R: A language and environment for statistical computing [Internet]. Vienna, Austria: R Foundation for Statistical Computing; 2021. Available from: https://www.R-project.org/

27. Kojima N, Klausner JD. Protective immunity after recovery from SARS-CoV-2 infection. Lancet Infect Dis. 2022 Jan 1;22(1):12–4.

28. Crotty S. Hybrid immunity. Science. 2021 Jun 25;372(6549):1392–3.

29. McGonagle DG. Health-care workers recovered from natural SARS-CoV-2 infection should be exempt from mandatory vaccination edicts. Lancet Rheumatol. 2022 Mar 1;4(3):e170.

30. Campus France. The health pass becomes the vaccine pass [Internet]. 2022 [cited 2022 Mar 25]. Available from: https://www.campusfrance.org/en/the-health-pass-becomes-the-vaccine-pass

31. The Government of the Hong Kong Special Adminstrative Region. Government adjusts vaccination requirements of Vaccine Pass [Internet]. [cited 2022 Mar 25]. Available from: https://www.info.gov.hk/gia/general/202203/20/P2022032000438.htm

32. German Missions USA. Coronavirus (COVID-19) [Internet]. [cited 2022 Mar 25]. Available from: https://www.germany.info/us-en/service/covid-19/2321562

33. Ferdinands JM. Waning 2-Dose and 3-Dose Effectiveness of mRNA Vaccines Against COVID-19–Associated Emergency Department and Urgent Care Encounters and Hospitalizations Among Adults During Periods of Delta and Omicron Variant Predominance — VISION Network, 10 States, August 2021–January 2022. MMWR Morb Mortal Wkly Rep [Internet]. 2022 [cited 2022 Apr 6];71. Available from: https://www.cdc.gov/mmwr/volumes/71/wr/mm7107e2.htm

34. Lamont N. STATE OF CONNECTICUT BY HIS EXCELLENCY NED LAMONT EXECUTIVE ORDER NO. 13G [Internet]. 2021. Available from: https://portal.ct.gov/-/media/Office-of-the-Governor/Executive-Orders/Lamont-Executive-Orders/Executive-Order-No-14B.pdf

35. Connecticut Government. Workplaces Subject to COVID-19 Vaccine Requirements [Internet]. http://CT.gov - Connecticut’s Official State Website. 2022 [cited 2022 Apr 7]. Available from: https://portal.ct.gov/Coronavirus/Covid-19-Knowledge-Base/Workplace-Vaccine-Requirements

