## Supplement for "Effectiveness of Primary and Booster COVID-19 mRNA Vaccination against Omicron Variant SARS-CoV-2 Infection in People with a Prior SARS-CoV-2 Infection"

**Supplement Table 1: Risk of SARS-CoV-2 Omicron Variant Infection among People Who Received Booster Vaccination Relative to Booster Eligible People*, According to Time after Receiving a Booster Vaccine Dose**

**Sensitivity Analyses:**

**Supplement Figure 1: Forest Plot of Vaccine Effectiveness from Matched Analysis, Stratified by the History of a Prior SARS-CoV-2 Infection**

**Supplement Figure 2: Number of Events Among Boosted Individual by Primary and Booster Dose Vaccine Brand**

**Supplement Figure 3: Forest Plot of Vaccine Effectiveness Exclusive of Test Among People with a Heterologous Booster Dose, Stratified by the History of a Prior SARS-CoV-2 Infection**

**Supplement Figure 4: Forest Plot of Vaccine Effectiveness Following the Inclusion of Tests Collected After Multiple Prior Positives, Stratified by the History of a Prior SARS-CoV-2 Infection**

**Supplement Figure 5: Forest Plot of Vaccine Effectiveness Exclusive of Discordant Thermo Fisher and Reflex Resulted Tests, Stratified by the History of a Prior SARS-CoV-2 Infection**

**Supplement Figure 6: Forest Plot of Vaccine Effectiveness Following the Inclusion of Inconclusive SGTF as Controls, Stratified by the History of a Prior SARS-CoV-2 Infection**

**Supplement Figure 7: Forest Plot of Vaccine Effectiveness Inclusive of all Controls, Stratified by the History of a Prior SARS-CoV-2 Infection**

**Supplement Figure 8: Forest Plot of Vaccine Effectiveness Following the Exclusion of Tests Among People with Prior Infections After a Vaccine Dose, Stratified by the History of a Prior SARS-CoV-2 Infection**

**Supplement Table 2: Risk of SARS-CoV-2 Omicron Variant Infection among People Who Received Booster Vaccination Relative to Booster Eligible People, According to History of a Prior SARS-CoV-2 Infection**

| **Supplement Table 1. Risk of SARS-CoV-2 Omicron Variant Infection among People Who Received Booster Vaccination Relative to Booster Eligible People^a^, According to Time after Receiving a Booster Vaccine Dose** | | | | |
| --- | --- | --- | --- | --- |
| **Vaccine status at testing** | **Case** | **Control** | **Odds ratio^b^** | **P-value^b^** |
| Boosted: 14-59 days after booster (3^rd^) dose | 267 | 5634 | - | - |
| Boosted: 60-89 days after booster (3^rd^) dose | 151 | 2037 | 1.15 (0.93, 1.43) | 0.197 |
| Boosted: 90+ days after booster (3^rd^) dose | 87 | 811 | 1.58 (1.21, 2.05) | 0.001 |
| ^a^ Limited to people without prior documented SARS-CoV-2 infections | | | | |
| ^b^ Adjusted for date of test, age (in years), sex, race/ethnicity, insurance, comorbidity (Charlson Score), SVI of zip code, presence of prior infection, municipality, and number of non-emergent visits during the year prior to vaccine rollout in Connecticut (December 2nd 2019 and December 1st 2020) | | | | |

**Sensitivity Analyses:**

*Matching (1:1 with replacement):* To allow for a single analytic sample from which to perform our analyses, we did not perform a match for our primary analysis. However, our fully adjusted (un-matched) model may suffer from positivity violations. To test if matching resulted in increased precision, we performed a 1:1 match with replacement on date of test (+/- 7 day), municipality, and presence of prior infections. The results of this analysis are below.

**Supplement Figure 1: Forest Plot of Vaccine Effectiveness from Matched Analysis, Stratified by the History of a Prior SARS-CoV-2 Infection**

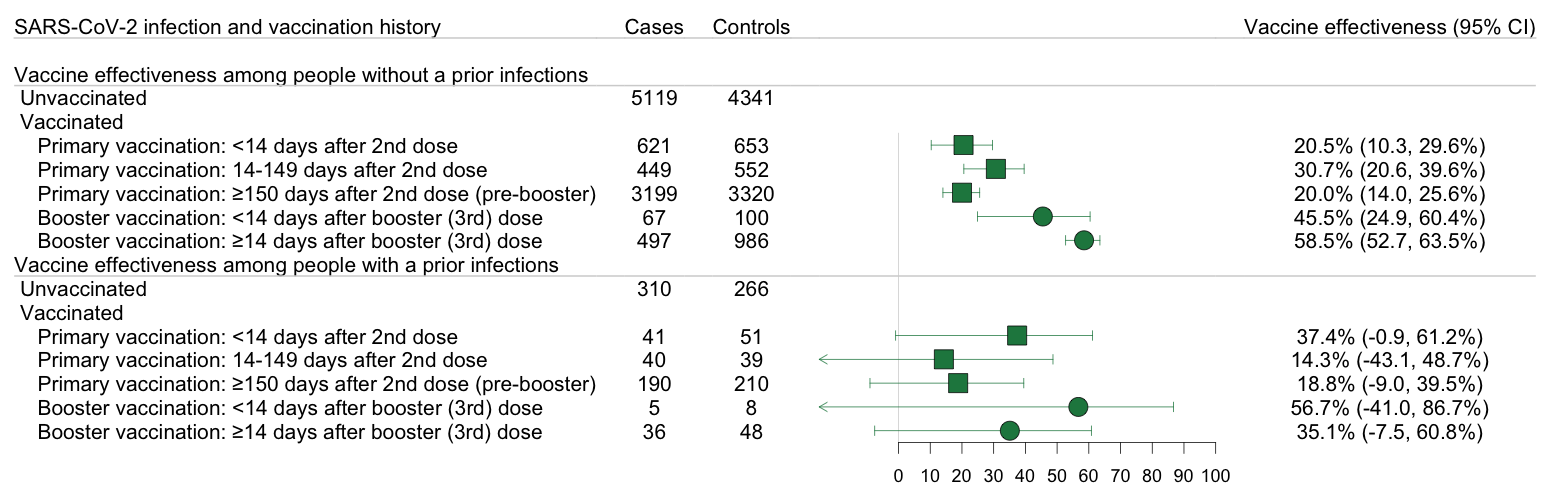

*Homologous booster doses:* We allowed for the inclusion of heterologous mRNA booster doses (ex. Moderna booster doses for people who received a primary Pfizer dose). While most people received homologous doses, <10% received a booster dose that did not align with their primary series. Four tests were collected among people who received a primary series mRNA and a booster J&J dose and were not considered in any analysis.

**Supplement Figure 2: Number of Events Among Boosted Individual by Primary and Booster Dose Vaccine Brand**

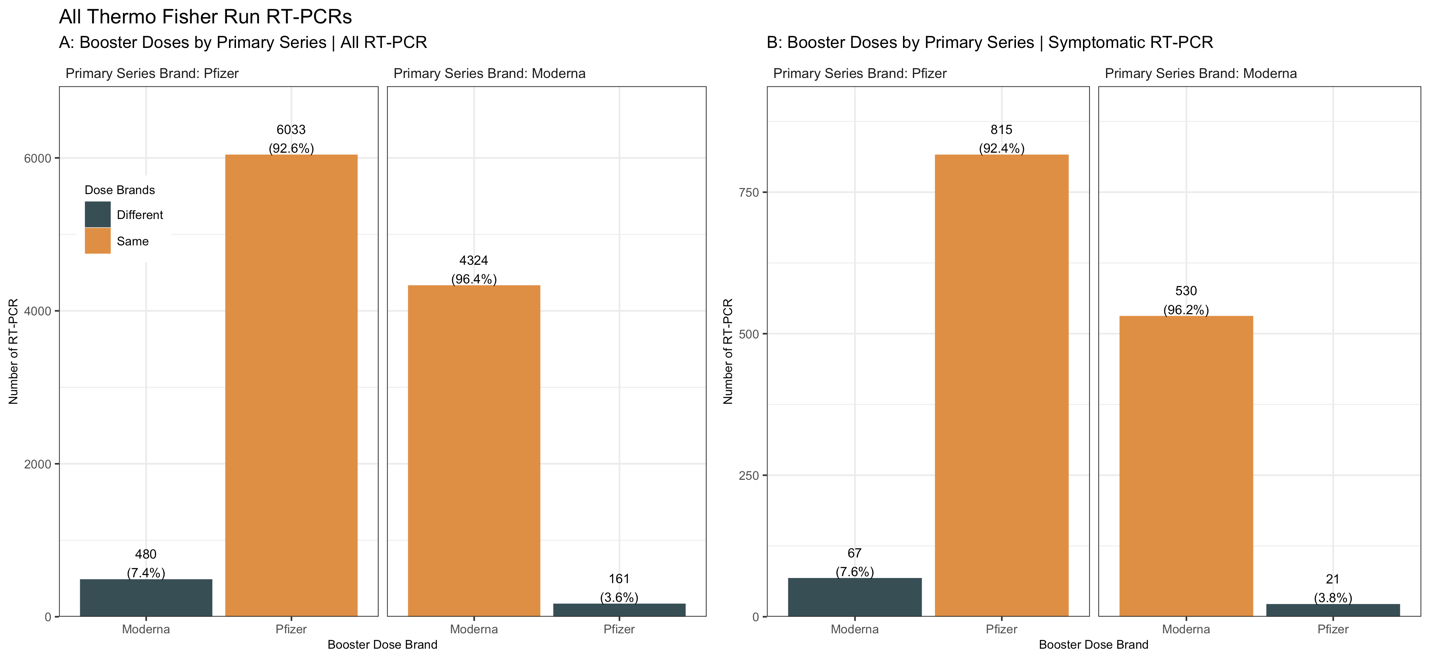

To test if discordant vaccine administration impacted our results, we restricted our sample to people who had homologous booster doses.

**Supplement Figure 3: Forest Plot of Vaccine Effectiveness Exclusive of Test Among People with a Heterologous Booster Dose, Stratified by the History of a Prior SARS-CoV-2 Infection**

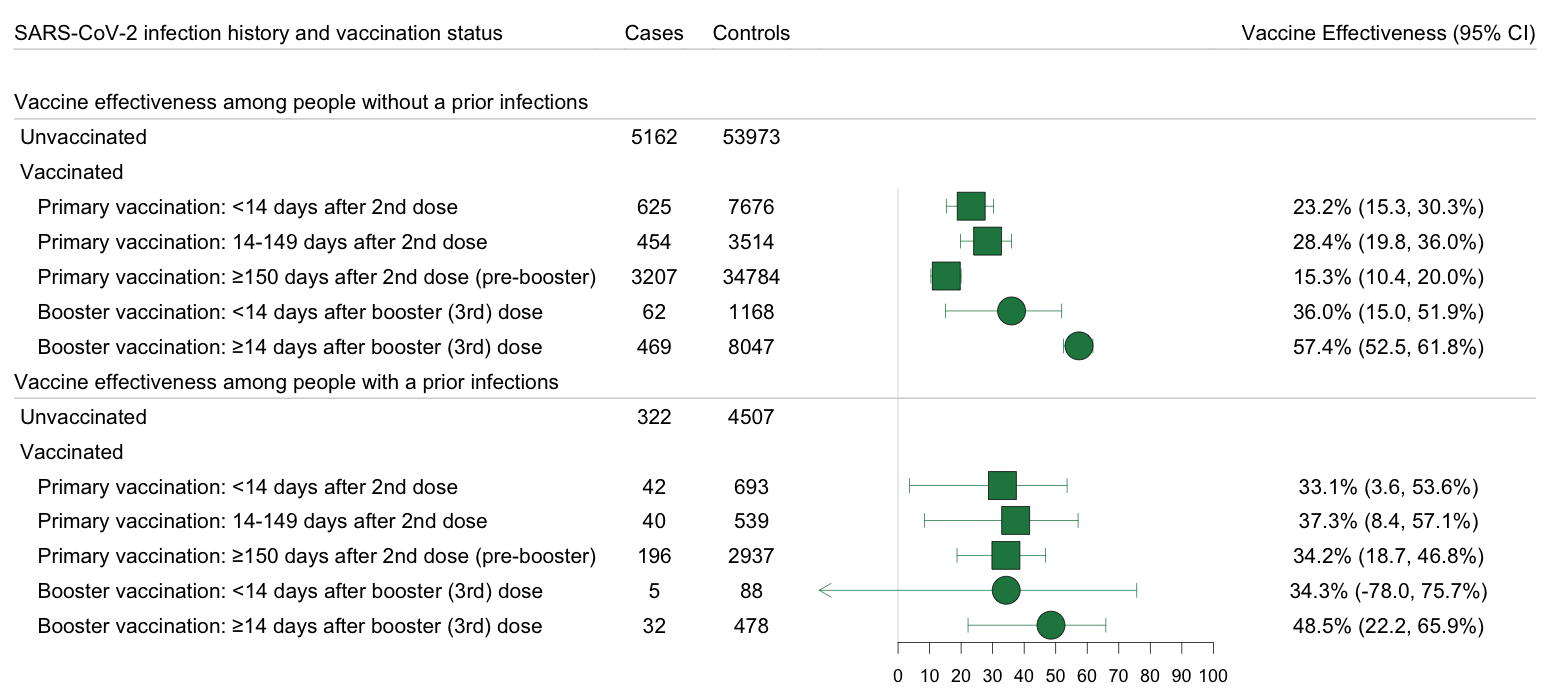

*Inclusion of tests collected among people with more than one prior infection:* In the primary analysis, we excluded tests collection after multiple prior positives. While this allowed us to estimate vaccine effectiveness among people with a prior infection, it was limited in its scope. Here we remove that restriction.

**Supplement Figure 4: Forest Plot of Vaccine Effectiveness Following the Inclusion of Tests Collected After Multiple Prior Positives, Stratified by the History of a Prior SARS-CoV-2 Infection**

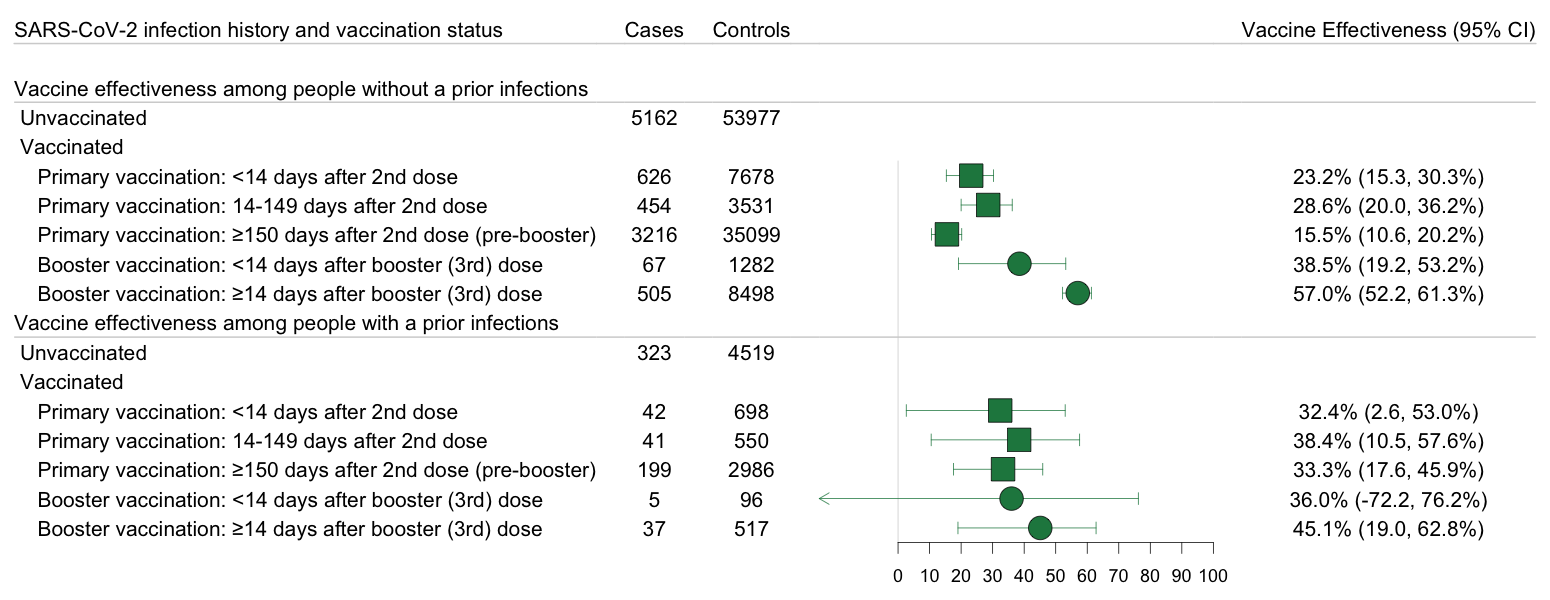

*Exclude Discordant Thermo Fisher and Reflex Tests:* In the primary analysis, we defined test positivity based on the results of the reflex test. Below are the vaccine effectiveness estimates when these 206 tests were dropped.

**Supplement Figure 5: Forest Plot of Vaccine Effectiveness Exclusive of Discordant Thermo Fisher and Reflex Resulted Tests, Stratified by the History of a Prior SARS-CoV-2 Infection**

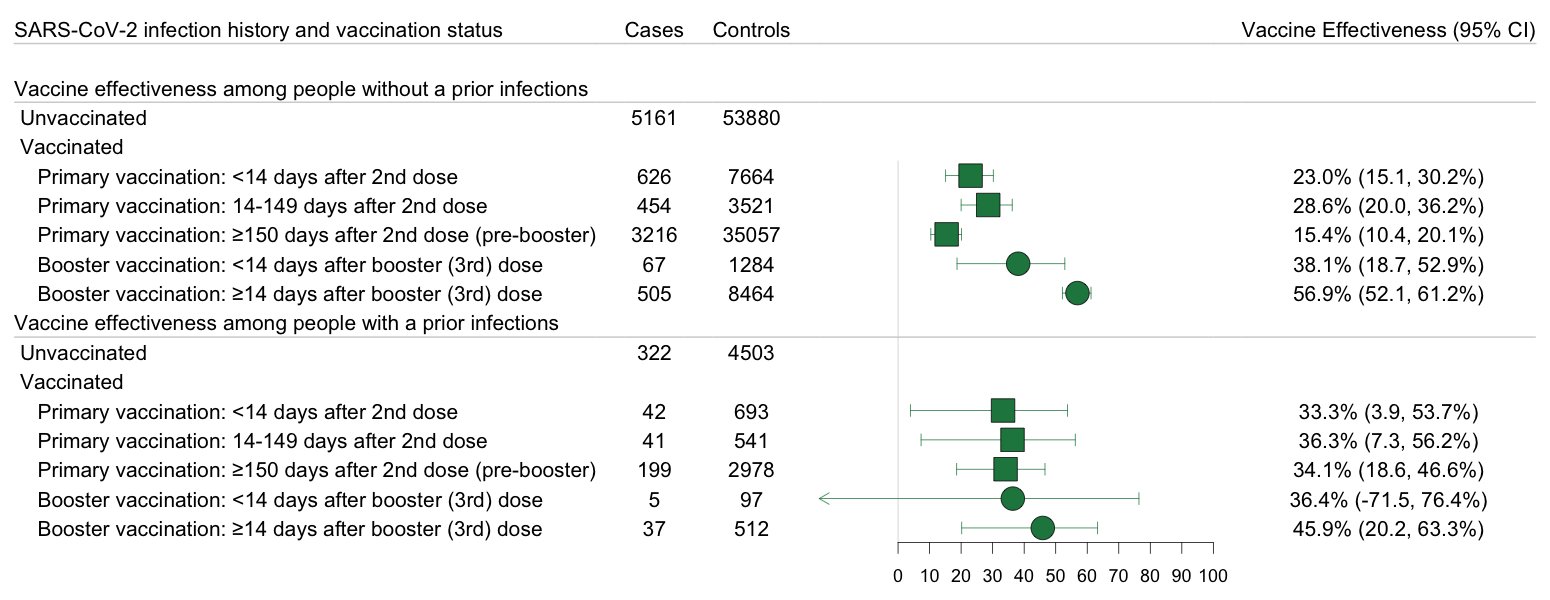

*Positive results with inconclusive SGTF listed as negative:* We defined presence of SGTF as [ORF1ab < 30] And [S Gene – ORF1ab >= 5] OR [ORF1ab < 30] And [S Gene >= 40]. If a sample was reported as positive but the ORF1ab read was > 30, SGTF was considered inconclusive. In the primary analysis, we excluded these positive results with inconclusive SGTF findings. This resulted in us having a clear variant designation for all positive RT-PCRs. Another way to handle such findings is to classify the sample as negative.^38^ Below are the vaccine effectiveness estimates when these tests were included as negatives.

**Supplement Figure 6: Forest Plot of Vaccine Effectiveness Following the Inclusion of Inconclusive SGTF as Controls, Stratified by the History of a Prior SARS-CoV-2 Infection**

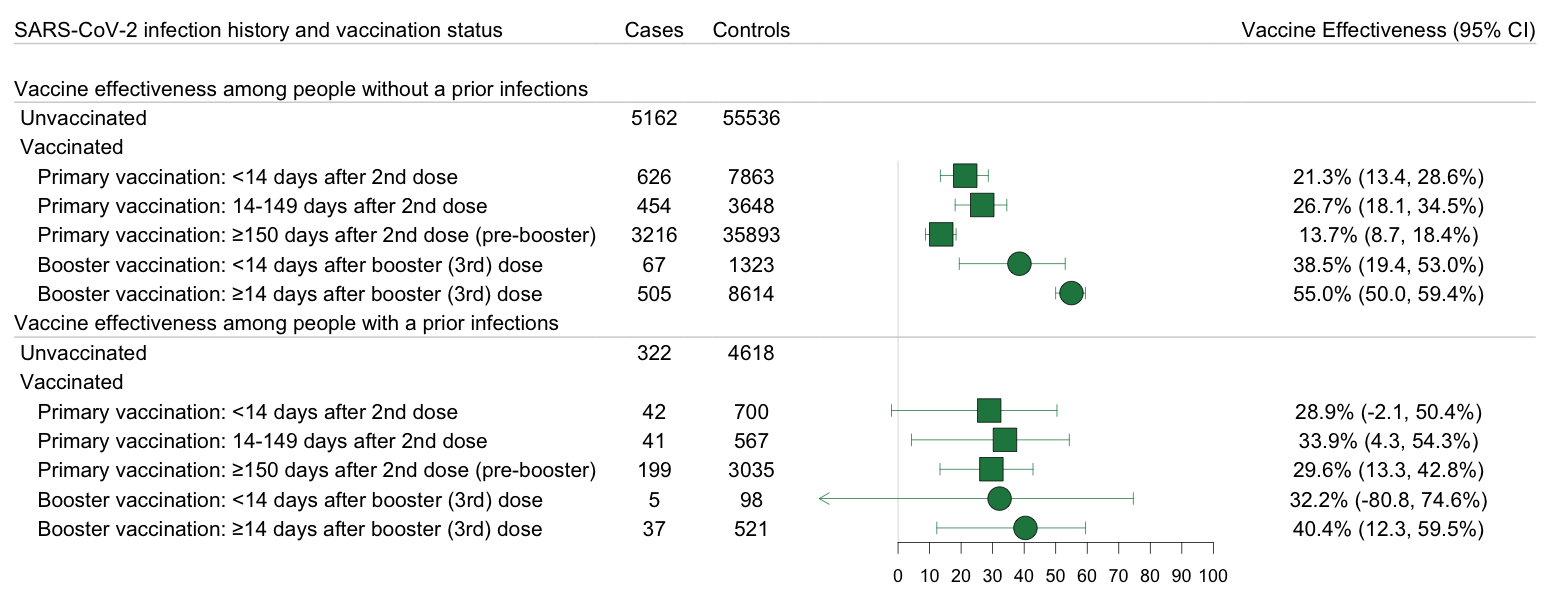

*All Controls:* To limit within person correlation within our controls, we limited the number of control events a person could contribute to three. Here we remove that restriction and allow for the inclusion of all controls.

**Supplement Figure 7: Forest Plot of Vaccine Effectiveness Inclusive of all Controls, Stratified by the History of a Prior SARS-CoV-2 Infection**

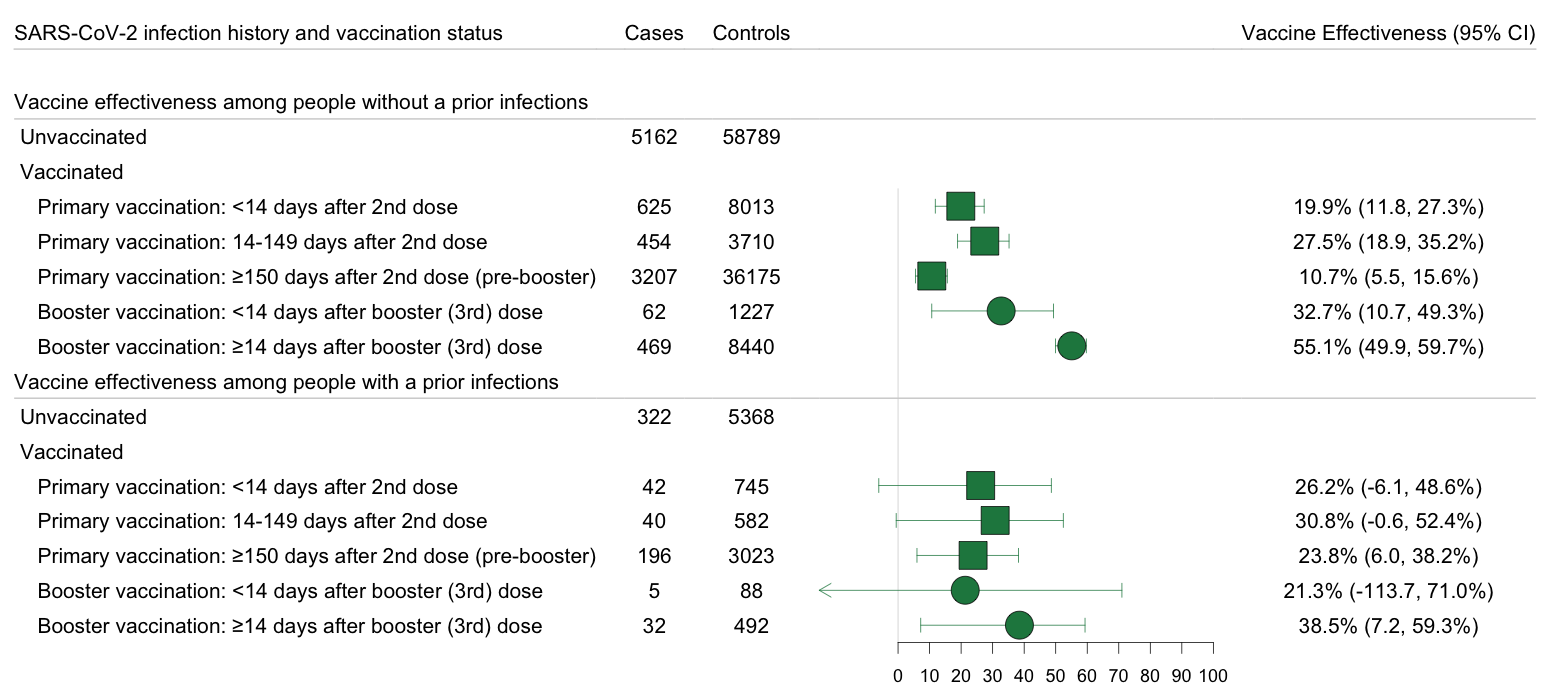

*Exclusion of tests collected among people with a prior infection after their first vaccination dose (prior breakthrough infection):* In the primary vaccine effectiveness analysis, we included breakthrough infections, or infections after a vaccination. This allowed us to give estimates of the level of protection vaccinated people with any prior infection have relative to people with only an infection. However, it did not directly evaluate the benefits of vaccination among people who have already been infected with SARS-CoV-2. Here, we excluded tests among people with prior infections that occurred after a vaccine dose.

**Supplement Figure 8: Forest Plot of Vaccine Effectiveness Following the Exclusion of Tests Among People with Prior Infections After a Vaccine Dose, Stratified by the History of a Prior SARS-CoV-2 Infection**

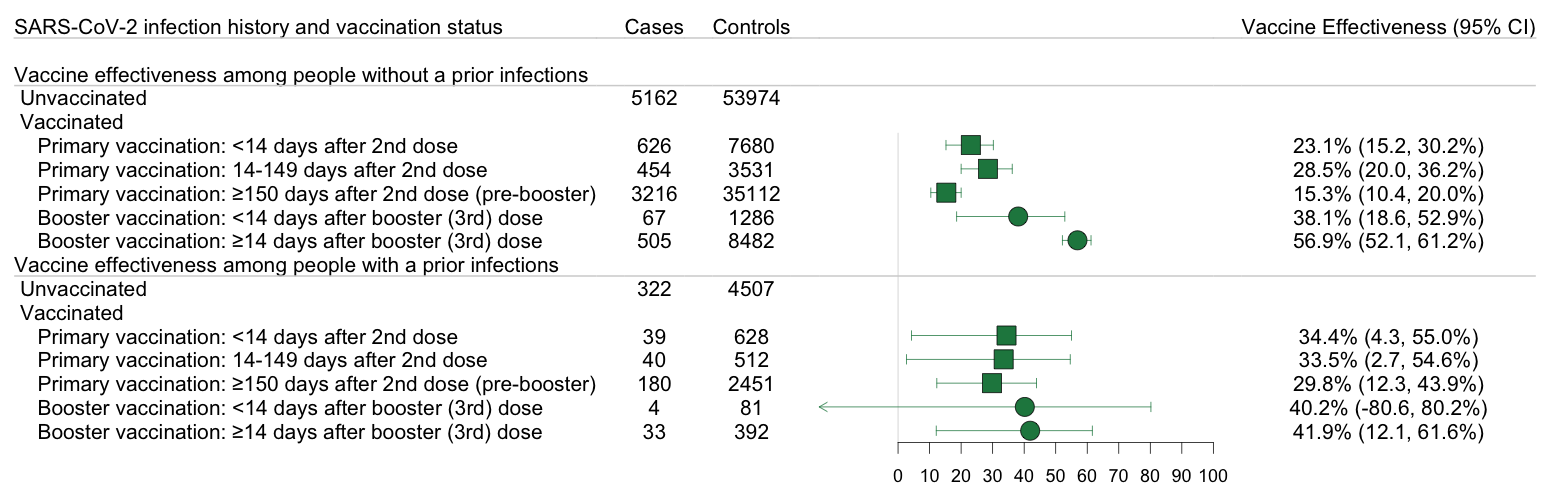

*Adjusting for time since referenced vaccine dose:* To examine the impact of booster doses among booster eligible people, we compared the odds of Omicron infection between boosted and booster eligible people stratified by presence of prior infection. In the primary analysis, the estimates came from a single model inclusive of all tests. Because of this, we were limited in our ability to adjust for time since primary series completion. Here, we account for time since primary series completion by restricting our sample to people who received at least two mRNA vaccine doses prior to testing. We, then adjusted for time since first primary series vaccine dose in a continuous manner using a natural spline with 3 knots.

| **Supplement Table 2: Risk of SARS-CoV-2 Omicron Variant Infection among People Who Received Booster Vaccination Relative to Booster Eligible People, According to History of a Prior SARS-CoV-2 Infection** | | | |
| --- | --- | --- | --- |
|  | **Omicron** | | |
| **Prior SARS-CoV-2 infection history and vaccination status** | **Cases** | **Controls** | **Odds Ratio^a^** |
| **With a prior infection^b^** |  |  |  |
| Booster eligible, ≥150 days after 2^nd^ dose (pre-booster dose)^c^ | 199 | 2982 | Reference |
| Boosted, ≥14 days after booster (3^rd^) dose ^d^ | 37 | 509 | 0.81 (0.53, 1.23) |
| **Without a prior infection^b^** |  |  |  |
| Booster eligible, ≥150 days after 2^nd^ dose (pre-booster dose)^c^ | 3216 | 35112 | Reference |
| Boosted. ≥14 days after booster (3^rd^) dose ^d^ | 505 | 8482 | 0.55 (0.49, 0.61) |
| ^a^ Adjusted for date of test, age, sex, race/ethnicity, insurance, Charlson Comorbidity Score, SVI of zip code, municipality, and number of non-emergent visits during the year prior to vaccine rollout in Connecticut (December 2nd 2019 and December 1st 2020) in all analyses and time between testing and last prior infection in analyses of people with prior infection. | | | |
| ^b^ Prior infection defined as a positive RT-PCR or rapid antigen test at least 90 days prior to included test | | | |
| ^c^ Booster eligible defined as primary series recipients who completed their primary series 150+ days prior to the test and were yet to receive a booster (third) dose; 150 days was selected as it reflects the CDC booster recommendations at the time of manuscript submission | | | |
| ^d^ Six tests were collected among people with a prior infection that occurred following their booster dose and were removed from this analysis | | | |
